# Clinical Development and Performance of the First To Know Syphilis Self-Test for Over-the-Counter Usage: a De Novo Rapid Test for Treponemal Antibody

**DOI:** 10.1101/2025.01.07.25320135

**Authors:** Kevin Clark, Sharon Rabine, Jianghong Qian, Diane Biddison, Sydney Spradlin, Vicki Thompson, Qinwei Shi, Jing Xu, Raji Pillai, Lu Zhang, Alicia Brown, Lisa Nibauer, Antony George Joyee, Jody D. Berry

## Abstract

The resurgence of syphilis in the USA and globally has hastened the need for widely available syphilis testing. Early detection of syphilis is crucial for avoiding serious clinical complications and preventing spread of infection. Self-tests enhance access to testing and promote timely treatment for positive cases. Herein we describe the performance of First to Know Syphilis Test (FTK), the first over the counter (OTC) treponemal test. A prospective multi-site clinical study with 1270 subjects was conducted, where subjects self-collected fingerstick capillary blood and self-tested without assistance. FTK test results were compared with a composite reference standard to evaluate diagnostic performance. The FTK test exhibited an overall sensitivity of 93.4% (95% CI, 87.0% to 96.8%) and specificity of 99.5% (95% CI, 98.9% - 99.8%) and accuracy of 99%. The overall agreement was 98.9%, with Cohens Kappa (ⱪ) value 0.93 (95% CI, 0.90 to 0.97) showing excellent agreement between the reference standard and FTK results. In addition, the test was validated in a panel of 125 clinically staged syphilis patient samples and the results showed 100% agreement in detecting anti-treponemal antibodies in all the samples. These data show excellent performance of the FTK Test demonstrating its utility in the screening and diagnosis of syphilis. The availability of this OTC test and its excellent performance at low prevalence, will have a profound impact on syphilis detection and prevention strategies and could reduce co morbidities such as HIV and other STI transmission.

## Introduction

Syphilis is a sexually transmitted infection (STI), caused by the spirochete bacterium, *Treponema pallidum*. It is transmitted by direct sexual contact or through vertical transmission from mother to child during pregnancy and can cause substantial morbidity and mortality. The number of syphilis cases in the U.S. is currently on the rise. According to the recent WHO report, infected cases have escalated globally by over 1 million in 2022, reaching a total of 8 million (1) that included 700,000 cases of congenital syphilis (2). The CDC reported that the cases increased by nearly 80% between 2018 and 2022 (from 115,000 to more than 207,000) (3). The rise in syphilis can be attributed to multiple factors including social and behavioral changes in this PrEP era, health disparities, particularly among sexual and gender minority populations, lack of timely testing and adequate treatment, intersections with the HIV and substance use epidemics, and increased maternal transmission leading to congenital syphilis infections (4). Syphilis is a chronic, multi-stage disease. The natural course of syphilis progresses through four stages including primary, secondary, latent and tertiary stages, with the primary and secondary stages being the most infectious. It must be treated in early stages to be cured. If left untreated, syphilis can lead to serious complications and permanent damage in the nervous and cardiovascular systems that can be life-threatening (5, 6, 7).

Syphilis during pregnancy, if untreated or treated late, can result in serious adverse birth outcomes including still births, neonatal deaths, prematurity, low birth weight, and congenitally infected infants (8, 9). Most people are often asymptomatic or do not notice symptoms, which makes screening a crucial step in diagnosis. There is no natural immunity to syphilis and the antibodies generated from past infection are insufficient to confer protection against re-infection.

The laboratory diagnosis of syphilis is complex and necessitates a combination of clinical and laboratory criteria to differentiate current or past infection, and absence of infection (10, 11). Culture for *T. pallidum* is cumbersome and is available only in selected research laboratories and there is no FDA-cleared PCR test for syphilis at present. Serological testing remains the mainstay for diagnosing syphilis infection. Serology is divided into two different tests: non-treponemal tests (NTTs) such as rapid plasma regain [RPR] test and Venereal Disease Research Laboratory (VDRL) and treponemal tests (TTs) such as *T. pallidum* Hemagglutination Assay (TPHA). While the TT is often positive for life, NTT titers decrease after treatment, therefore the current TTs and NTTs have their limitations, hampering syphilis control and prevention efforts in real-world settings (9). A single serologic NTT or TT is not sufficient for diagnosis and therefore a combination of positive TT and reactive NTT is required for the diagnosis of syphilis infection. In the past, syphilis screening mainly followed a traditional testing algorithm, starting with NTT and the positive results require further confirmation through more specific TTs (9, 10). With increased availability of various treponemal-specific immunoassays including enzyme immunoassays (EIAs), chemiluminescence immunoassays (CIAs) etc., recently, many laboratories are increasingly adopting a ‘reverse sequence screening’ algorithm which begins the screening with a TT and the reactivity is confirmed with an NTT. When the NTT is non-reactive, a second but different treponemal test is performed to determine if the first treponemal test was a false positive (12). Regardless of which algorithm is used, it is important to consider the sensitivity and specificity of these assays in clinically characterized sera, stratified by stage of syphilis (13) and all screening results must be correlated with clinical diagnosis covering patients’ symptoms, previous history and sexual risk factors to make an accurate diagnosis (14).

Frequent syphilis testing for timely and accurate diagnosis of active infections for appropriate clinical management is a key strategy to effectively prevent disease transmission (7). This could be achieved with rapid point-of-care tests which are easy to perform, and inexpensive so that repeated testing is practical, and test results are available immediately. Rapid over the counter (OTC) tests have become entrenched in society since the COVID-19 pandemic. There is a broad acceptance of being able to self-test at home. An in-home test for syphilis has the potential to greatly impact public health by improving access to timely detection and treatment.

Herein we describe the clinical performance of the first over-the-counter (OTC) treponemal rapid test. The First To Know^®^ Syphilis Test (NOWDiagnostics Inc.) is the first at-home, OTC test for syphilis, having received FDA marketing authorization under a de novo classification pathway in August 2024. It is a rapid buffer-less lateral flow immunoassay test device that detects *T. pallidum* antibodies in human whole blood (capillary) from individuals suspected of having a syphilis infection. Here, we report the results from a clinical study that evaluated the test performance of the First To Know^®^ Syphilis Test.

## Materials and methods

The First To Know^®^ Syphilis Test is a simple buffer-less lateral flow visually-read test device that detects antibodies to syphilis antigens and requires a fingerstick and whole blood to run the test in as little as 15 minutes. There is no need for a swab nor a stand to run this test. Briefly, a drop of human capillary whole blood is added to the test device cassette, where the blood is drawn into the fill zone. As the sample flows into the porous test strip, syphilis-specific antibody in the sample binds at the test band location. The appearance of a visible test band indicates the sample contains a detectable level of anti-treponemal antibody. The internal control and the appearance of the control band line assures that the sample is applied correctly, and the test components are working properly. The test is a chromatographic immunoassay that delivers results in 15 minutes.

### Study subjects, sample collection, and Syphilis testing

To evaluate the performance of the First To Know^®^ Syphilis Test, a prospective clinical study was conducted in six geographically diverse sites across the US (the sites were in Tempe, AZ; Las Vegas, NV; Los Angeles, CA; Spokane, WA; West Orange, NJ and North Miami, FL). Individuals who met the inclusion criteria were consecutively included in the study. The sample size for the test performance evaluation was determined based on the FDA guidance documents. For the clinical study, subjects from two cohorts were enrolled (September 1, 2021, through October 17, 2023); One cohort of sexually active persons (18-64 years old) and another cohort of expectant mothers ( ≥18 years old). The enrolled subjects self-collected capillary blood from a finger stick and executed self-testing according to the instructions for use with the First To Know^®^ Syphilis Test. The clinical study protocol and the informed consent form were approved by an IRB. If the test results were incomplete or not performed or lab results were missing for any samples, they were excluded from the analysis. Of the 1345 subjects enrolled, samples from 75 subjects had invalid results and were excluded. A total of 1270 subjects from the two cohorts who obtained a valid result were included (1011 samples from the sexually active cohort and 259 samples from the expectant mothers cohort combined for the final evaluation) to evaluate the diagnostic performance of the First To Know^®^ Syphilis Test. There were no adverse events in the performance study.

### Composite reference result

Venous whole blood samples were also collected from the subjects, plasma and serum samples prepared from whole blood were sent and tested by a reference lab using FDA-cleared comparator tests. The comparator tests included two treponemal tests (BioPlex 2200 Syphilis Total and SERODIA-TP-PA Test) and one non-treponemal test (Wampole Impact RPR Test). The reference diagnosis of ‘true positive’ for syphilis was established if at least two of the three tests were positive, and this composite reference standard was used for comparison with the First To Know^®^ Syphilis Test to evaluate test sensitivity and specificity. The participants did not have reference standard test results at the time of self-testing with the First To Know^®^ Syphilis Test. The Standards for Reporting of Diagnostic Accuracy Studies (STARD) reporting guidelines were followed to ensure the completeness and transparency of reporting diagnostic accuracy studies (15).

### Clinically staged syphilis patient samples and testing with First To Know^®^ Syphilis Test

In addition to the clinical study evaluation, we validated the diagnostic accuracy of the First To Know^®^ Syphilis Test in a panel of characterized serum samples of known positive clinically staged patients obtained from the CDC. The panel included serum samples from patients with primary syphilis (n=25), secondary syphilis (n=56), early latent syphilis (n=16), and late latent syphilis (n=28).

### Statistical analysis

Statistical analysis was performed using GraphPad Prism 9 (GraphPad Software, Inc.) The performance of the First To Know^®^ Syphilis Test was evaluated based on comparison with composite reference standard using a dichotomous approach, two by two crosstab analysis for the calculation of sensitivity (positive percent agreement [PPA]), specificity (negative percent agreement [NPA]), positive predictive value (PPV), and negative predictive value (NPV) and accuracy. The concordance analysis for diagnostic performance was determined using the Cohen’s kappa (κ) value according to the criteria proposed by Landis & Koch, e.g. values more than 0.8 were considered as “almost perfect agreement” (16).

## Results

Samples from a total of 1270 study participants (that consisted of two cohorts including a sexually active cohort and a pregnant women cohort) were subjected to this evaluation study for the First To Know^®^ Syphilis Test. The overall positivity for syphilis in this study was determined to be 8.3% (106/1270) based on the composite reference standard. The First To Know^®^ test showed positivity in 99 out of the 106 positive samples. Among the 1164 negative samples, 1158 samples were negative by the First To Know^®^ test. Based on the composite reference test results, the First To Know^®^ test yielded an overall sensitivity of 93.4% (95% CI, 87.0% - 96.8%) and specificity of 99.5% (95% CI, 98.9% - 99.8%) (Table 1). The test demonstrated an accuracy of 99% in this evaluation. The overall agreement between the reference standard and the First To Know^®^ test was 98.9%, with Cohen’s Kappa (ⱪ) value 0.93 (95% CI, 0.90–0.97) showing within the scale of ‘almost perfect agreement’ (Kappa between 0.81 and 1.00).

**Figure 1.**
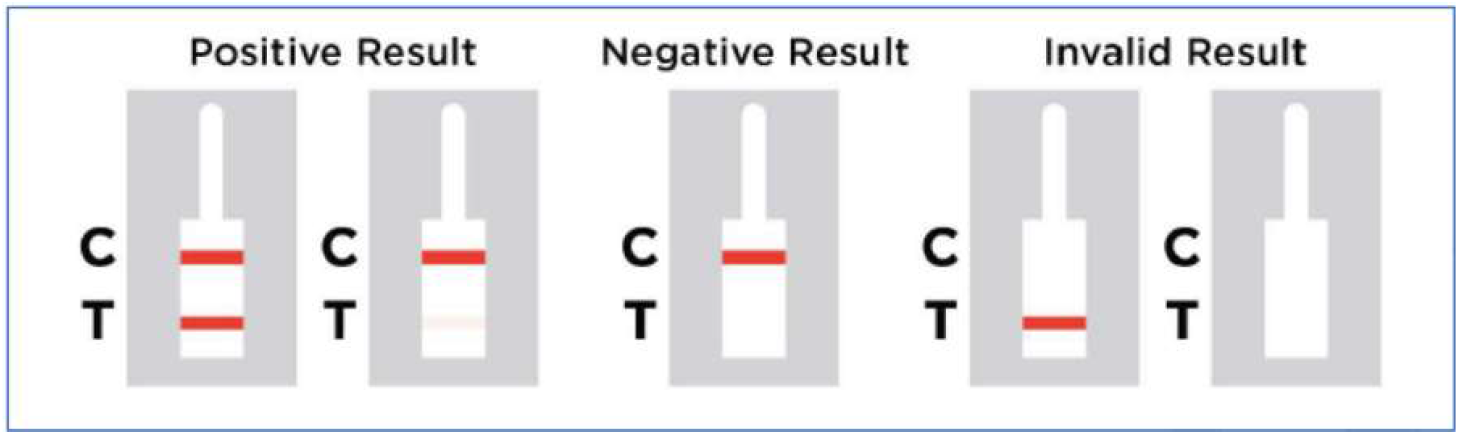
First To Know^®^ Syphilis test device showing visually read test results.

**Table 1.** Evaluation of performance characteristics of First To Know^®^ Syphilis test compared to the composite reference as authorized by the FDA for OTC.

| First To Know<sup>®</sup><br>assay | Reference (composite) |  |  |
| --- | --- | --- | --- |
|  | Positive | Negative | Subtotal |
| Positive | 99 | 6 | 105 |
| Negative | 7 | 1158 | 1165 |
| Subtotal | 106 | 1164 | 1270 |
| Sensitivity/PPA | 93.4% (95% CI, 87.0% - 96.8%) |  |  |
| Specificity/NPA | 99.5% (95% CI, 98.9% - 99.8%) |  |  |
| Positive Predictive Value (PPV) | 94.3% (95% CI, 88.1% - 97.4%) |  |  |
| Negative Predictive Value (NPV) | 99.4% (95% CI, 98.8% - 99.7 %) |  |  |
| Accuracy | 99.0% (95% CI, 98.3% - 99.5%) |  |  |

\* For reference standard, positive and negative results were determined by a composite algorithm as described in the methods.

When the cohorts in the study population were analyzed separately, the sensitivity and specificity of the First To Know^®^ test for the sexually active cohort group samples (n=1011), were 94.9% (95% CI, 88.5% - 98.3%) and 99.6% (95% CI, 98.9% - 99.9%) respectively, while in the pregnant women cohort (n=259), the sensitivity and specificity of the test were 75.0% (95% CI, 40.9% - 92.9%) and 99.2% (95% CI, 97.1% - 99.8%) respectively. The First To Know^®^ test results observed with different cohorts are shown in Table 2.

**Table 2.** Study cohorts and test performance.

| Study cohorts<br>Total (n=1270) | **First To Know® Syphilis Test** |  |
| --- | --- | --- |
|  | Sensitivity/PPA<br>Test +/Ref + (% , 95% CI) | Specificity/NPA<br>Test -/Ref - (% , 95% CI) |
| Sexually active cohort (n=1011) | 93/98 (94.9%, 95% CI, 88.6% - 97.8%) | 909/913 (99.6%, 95% CI, 98.9% - 99.8%) |
| Pregnant cohort (n=259) | 6/8 (75.0%, 95% CI, 40.9%- 92.9%) | 249/251 (99.2%, 95% CI, 97.1% - 99.8%) |

In addition to the above, we also evaluated the First To Know^®^ Syphilis Test in a set of 125 characterized serum samples from known positive clinically staged patients. The First To Know^®^ Syphilis Test showed 100% sensitivity in detecting anti-treponemal antibodies in all the samples tested, having absolute agreement with reference standard. The breakdown of different clinically staged groups and test positivity are shown in Table 3.

**Table 3.** First To Know^®^ Syphilis testing in Clinically staged syphilis patients (n=125).

| **Clinical stage of Syphilis**<br>**Samples (n=125)** | **Number Tested** | **Test positivity**<br>**Number (%)** |
| --- | --- | --- |
| Primary Syphilis | 25 | 25 (100%) |
| Secondary Syphilis | 56 | 56 (100%) |
| Early Latent Syphilis | 16 | 16 (100%) |
| Late Latent Syphilis | 28 | 28 (100%) |

## Discussion

Syphilis is a preventable and curable bacterial sexually transmitted infection but if left undiagnosed and untreated, it can result in serious downstream health outcomes. Further, there is a higher risk of acquiring HIV and other STIs (17, 18). Therefore, rapid and accurate detection of syphilis is indispensable to ensure treatment and to control the transmission of the disease. In the present study, we evaluated the diagnostic performance of the First To Know Syphilis^®^ Test, in a large real world lateral flow study. The findings of present evaluation based on self-testing by the study subjects showed excellent overall clinical performance of the First To Know^®^ test (93.4.% sensitivity, 99.5% specificity and 99% accuracy) compared with the reference method. This also meets the WHO recommended criteria of minimum of 85% sensitivity and 95% specificity for syphilis rapid tests (19). The sensitivities of commercially available POC tests are in the range of 76% - 98%, with 90% to 99% specificities in varied settings. (20-25). In addition to the clinical performance study, we validated the diagnostic accuracy of the First To Know^®^ test in a panel of characterized samples from clinically staged syphilis patients. Our results showed that the First To Know^®^ test showed 100% sensitivity in detecting anti-treponemal antibodies in all the tested samples in the panel demonstrating diagnostic efficacy. These results suggest that the First To Know^®^ test has the potential to aid accurate syphilis diagnosis in high-risk patient populations with specific clinical stages, however larger and more detailed studies are needed to confirm the present finding.

The current resurgence of syphilis in the US and the alarming spread of infections underscores the critical need for rapid diagnosis and containment (2, 3, 26). Currently, syphilis incidence is increasing more rapidly among men who have sex with men (MSM) than any other U.S. subpopulation (27, 28). Consistent with literature, we observed that syphilis test positivity in MSM was significantly higher than that of non-MSM (32.8% vs 6.9%, p<0.001) (data not shown). In addition, there is a dramatic increase in congenital syphilis over the 4-year period from 2018 to 2022 (3) which clearly necessitates early detection. Increasing the coverage and frequency of syphilis testing are crucial elements of any control effort to allow earlier detection and treatment, interrupting transmission which aids to control the spread of syphilis. Specifically, focusing testing on high-risk populations like men who have sex with men (MSM), female sex workers and routine syphilis testing during pregnancy can significantly impact syphilis control. The de novo FDA marketing authorization and availability of the First To Know^®^ Syphilis Test represent a key advancement in the landscape of rapid diagnostics for syphilis that would enable increased access to testing and early diagnosis and will support the universal effort in preventing transmission of infections. Although POC tests were available prior to the First To Know^®^ marketing authorization, currently, the First To Know^®^ Syphilis Test is the only at-home, OTC test for syphilis. It should be noted that positive test results from this test should be followed by additional laboratory testing through a health care provider to confirm the diagnosis. Nevertheless, it serves best as a screening tool to warn individuals that they may have an identifiable infection.

While syphilis rates have risen considerably in high-risk populations, its prevalence is still relatively low in the overall population. The setting of our clinical performance study reflects a relatively low-prevalence environment, resembling more of a community-based setting. In an analysis comparing First To Know^®^ test with two other U.S. commercial POC tests, we observed that the PPVs of the First To Know^®^ test remained the highest and did not show dramatic decrease with a decrease in prevalence rates compared to the other two POC rapid tests. The NPVs showed no statistical difference between the tests (supplemental, Figure S1). This suggests that First To Know^®^ test could accurately identify and refer positives from testing to further care (ruling in those at risk) and also could identify those who are negative. Overall, these observations and above findings indicate the reliability of the First To Know^®^ test in early detection and management of the disease that can lead to better population health outcomes. Real world studies in different prevalence settings/populations are further needed to solidify the diagnostic efficacy and clinical utility of the test.

The availability of an over-the-counter test with at-home results like the First To Know^®^ Syphilis Test has important implications that extend beyond testing for convenience. By being readily available, it could increase the initial volume of screening, which is the first step towards diagnosing syphilis, which is crucial for early treatment and prevention of serious complications. Home based self-testing for STIs like syphilis provides several benefits over conventional clinic/laboratory-based testing. In addition to the simplicity in performing the test, it provides privacy, without fear of judgement and therefore offers significant advantages over currently available POC tests. Self-testing at home can be a beneficial strategy to reach more at-risk individuals who are reluctant or otherwise may never seek testing. In fact, it has been proven effective in the case of HIV control programs and helped to counter barriers for testing those at high-risk who otherwise rarely get tested (29, 30). The recent CDC randomized clinical trial called Evaluation of Rapid HIV Self-testing Among MSM Project (eSTAMP) study showed that self-testing increased the uptake and frequency in testing and resulted in people seeking care or additional testing after obtaining a positive result (31). These programs and other studies in the U.S. exemplify the high level of acceptability and effectiveness of home testing for STIs (29). With the availability of an OTC test like the First To Know^®^ test, it is feasible to implement similar programs for syphilis which could help control spread of infection. In this regard, the First To Know^®^ Syphilis Test’s ease of use with a simple fingerstick blood micro sampling and short time to obtain results with high sensitivity and specificity can be very advantageous for self-testing and to facilitate early diagnosis. Our data showed that the First To Know^®^ test is robust for self-testing in different settings; the usability studies demonstrated that the error rate during self-testing with First To Know^®^ was very minimal (data not shown) underscoring the ease of use and suitability for testing by lay users.

There are some limitations to our study; the study population represents a low prevalent setting, and the performance in high prevalence groups was not evaluated here. Therefore, the results from the current study might not reflect its performance in higher prevalence settings thus possibly underestimating the test performance. While our study included a pregnant cohort, the sample size was small. Considering the importance of syphilis testing in pregnant women, it is important to have further detailed clinical studies in pregnant women. In addition, future research involving larger, clinically stratified patient populations will be important to strengthen our findings.

In conclusion, the present study observations showed an excellent overall diagnostic performance of the First To Know^®^ Syphilis Test demonstrating its utility as a valuable tool in the screening and diagnosis of syphilis. As the incidence of syphilis continues to rise in the United States and worldwide, the availability of this OTC test can have a profound impact on syphilis detection and prevention strategies and could support reduction of co-morbidities such as HIV and other STI transmission.

## Author Contributions

Conceptualization: KC, VT; Data curation: SR, DB, VT, LZ; Formal Analysis: DB, AGJ, LN, RP; Funding Acquisition: KC; Experimentation: KC, JQ, SS, VT, LZ, AB, JX, QS; Methodology: JQ, KC,QS, JX, SS, AB; Supervision: KC, VT; Validation: DB, SR; Resources: DB, AB; Project Administration: SR, RP; Writing Original Draft Preparation: AGJ, JDB; Writing Review & Editing: AGJ, JDB, KC, LN. All authors have read and agreed to the published version of the manuscript.

## Ethics Statement

This study was conducted in accordance with the ethical standards of the Helsinki Declaration. IRB approval for the multi-site study was obtained from Advarra IRB #Pro00032035. The initial approval was on Dec 9, 2019, and latest approval was on July 25, 2023. Informed consent was obtained from all study subjects, and all the participant data were de-identified in the database. Clinicaltrials.gov registration number NCT05063344

## Data Availability Statement

All data supporting the findings of this research study are described in the manuscript and relevant details are available upon reasonable request. Due to the involvement of human subjects and nature of the STI study, the datasets are not shared in public repositories due to the limitations on ethical and privacy concerns.

## Acknowledgments

We thank Dave Kern, regulatory consultant, for his support for this work. We greatly thank Robert Weigle, Joshua Bafford, Thomas Grifa, Susan Ward, and Todd Grice for their continued support and technical inputs into the program and the manuscript. We thank Jing Shi, PhD for statistical guidance and inputs, Deonna Robertson for technical assistance and Todd Simpson for graphic arts support.

## Supplemental

**Figure S1.**
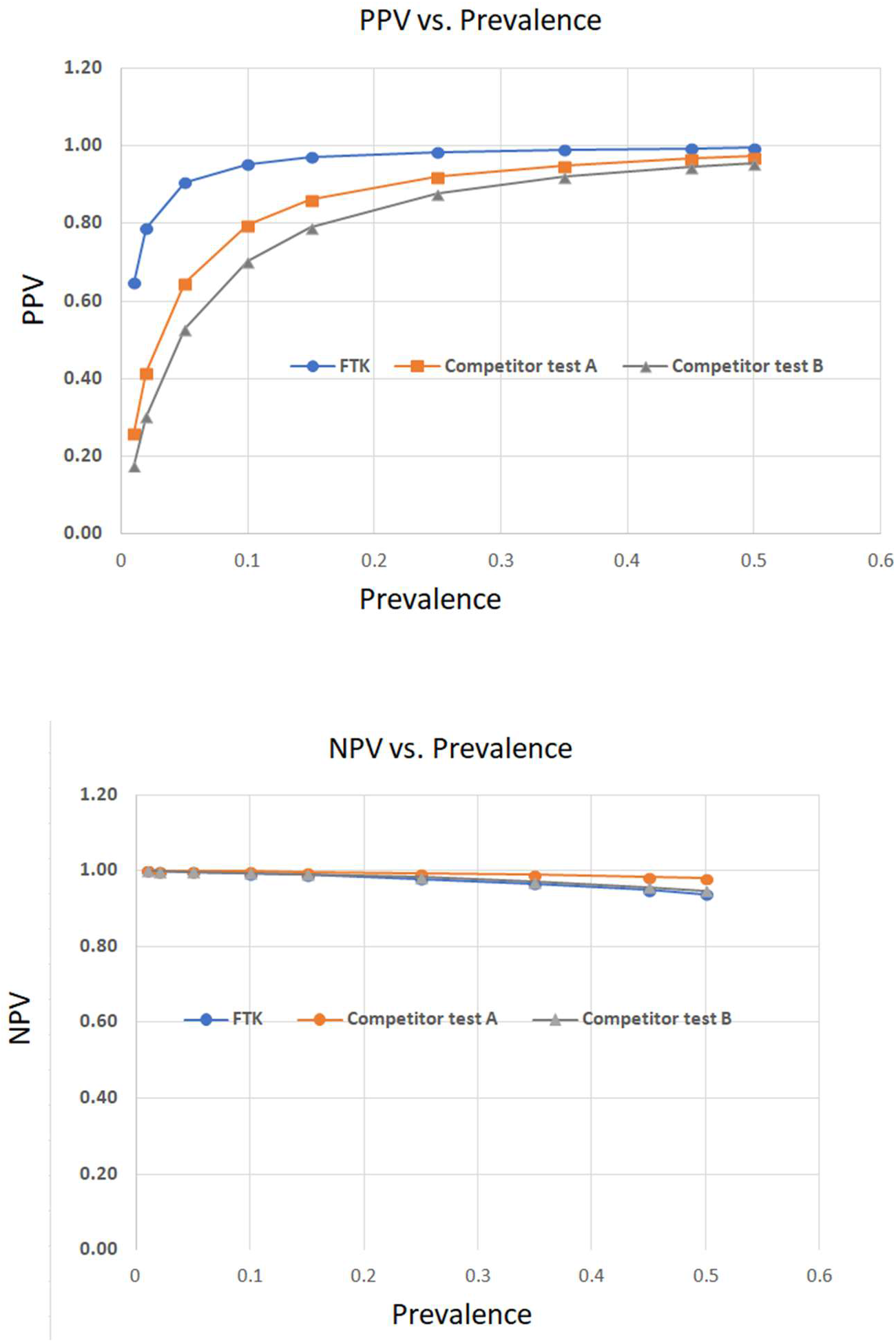
Comparative analysis of First To Know^**®**^ Syphilis Test with other commercial tests on PPV and NPV with different prevalence settings based on sensitivities and specificities. *PPV and NPVs were based on sensitivities and specificities of different tests. FTK; First To Know^**®**^ Syphilis Test

